# Differential association of step count with depressive and anxiety symptoms in older adults at risk of dementia

**DOI:** 10.64898/2026.07.12.26357854

**Authors:** Yolanda Lau, Harisd Phannarus, Claudia Cooper, Zuzana Walker, Harriet Demnitz-King, Natalie L Marchant

**Affiliations:** Division of Psychiatry, University College London, London, UK; Centre for Psychiatry and Mental Health, Wolfson Institute of Population Health, Queen Mary University of London, London, UK; Department of Preventive and Social Medicine, Faculty of Medicine Siriraj Hospital, Mahidol University, Thailand; Essex Partnership University NHS Foundation Trust, Essex, UK

## Abstract

**Background:** Depressive and anxiety symptoms are prevalent among older adults and associated with increased dementia risk. In healthy older adults, higher step counts are associated with fewer depressive and anxiety symptoms; whether this holds in individuals with cognitive concerns (subjective cognitive decline [SCD] or mild cognitive impairment [MCI]) is unknown. In a randomised controlled trial, the 12-month APPLE-Tree group psychosocial lifestyle intervention produced small cognitive improvements but no change in step count.

**Objective:** To test whether step count was associated with depressive and anxiety symptoms (cross-sectionally and over 24 months), and whether APPLE-Tree increased step count in participants with clinical anxiety or depression.

**Methods:** We examined cross-sectional and longitudinal (12- and 24-month) associations between step count (two-week average from wrist-worn wearables) and depressive and anxiety symptoms (Hospital Anxiety and Depression Scale) using adjusted linear regressions, with a mediation analysis of self-perceived mobility. We also tested whether the intervention increased step count in those with baseline clinical anxiety or depression.

**Findings:** We included 629 of 746 trial participants at baseline, of whom 376 contributed 12-month and 215 24-month data. At baseline, higher step counts were associated with fewer depressive symptoms (β = −0.11, 95% CI −0.17 to −0.05, p < 0.001) but, counter to our hypothesis, more anxiety symptoms (β = 0.12, 95% CI 0.06 to 0.19, p = 0.003). Over two years, change in step count was not associated with change in depressive or anxiety symptoms (all p ≥ 0.12). Self-perceived mobility problems mediated the association between step count and depressive but not anxiety symptoms. The intervention did not change step count in those with clinical anxiety or depression.

**Conclusions:** This provides the first evidence in older adults with cognitive concerns that higher step counts are associated with fewer depressive but more anxiety symptoms. This may reflect heterogeneity of a population that includes those with prodromal dementia and cognitive health anxiety. Step count did not predict symptoms over time.

**Clinical implication:** Step count may help distinguish anxiety and depressive symptoms in people presenting with cognitive concerns, or underlying reasons for cognitive concerns among those with functional cognitive disorders.

**KEY MESSAGES:** **What is already known on this topic** – *summarise the state of scientific knowledge on this subject before you did your study and why this study needed to be done*

Physical inactivity, depressive symptoms, and anxiety symptoms are linked to negative health outcomes and increased dementia risk. Existing research has shown that physical activity (e.g., walking) is associated with lower depressive and anxiety symptoms in healthy adults, and physical activity interventions are increasingly being employed to reduce these symptoms. Such interventions could offer benefit for individuals at risk of dementia; however, it remains unknown whether the relationship between physical activity and depressive and anxiety symptoms is the same in people with cognitive concerns, who may struggle to engage in planned physical activities.

**What this study adds** – *summarise what we now know as a result of this study that we did not know before*

This study extends existing research by showing that higher step counts were associated with fewer depressive symptoms in older adults at risk of developing dementia (i.e., with subjective cognitive decline [SCD] or mild cognitive impairment [MCI]). However, contrary to existing research, higher step counts were associated with more anxiety symptoms in this population. Self-perceived mobility problems fully mediated the association between step count and depressive symptoms, but did not mediate the association with anxiety symptoms, suggesting that the mechanisms linking step count with depressive and anxiety symptoms differ in older adults at risk of dementia. In longitudinal analyses, neither the intervention nor change in step count was associated with change in symptoms, indicating that step count may be a correlate of symptoms rather than a modifiable target in this population.

**How this study might affect research, practice or policy** – *summarise the implications of this study*

Walking interventions may be most appropriate for older adults with SCD or MCI experiencing depressive symptoms. Several studies have questioned whether among individuals with obsessional anxiety symptoms who have eating disorders, fitness trackers may reinforce worries. In people presenting with cognitive concerns without dementia, anxiety symptoms will have heterogenous aetiologies: worries about memory loss, as well as health anxieties that include fears of getting dementia. It is important to reflect on the impact of generic messaging on different groups, and perhaps to consider in further research the possibility that encouraging use of exercise trackers may compound these anxieties for a minority. For policymakers and practitioners, this underlines the importance of tailoring dementia prevention advice.

## BACKGROUND

Dementia is a neurodegenerative condition that affects millions of individuals worldwide. As of 2019, 57.4 million individuals were living with dementia globally, with this number estimated to triple by 2050^1^. Despite recent advancements, no disease-modifying treatments are widely available, highlighting the importance of prevention and early intervention. Research has increasingly focused on identifying modifiable factors that may reduce dementia risk or delay its onset^2^, particularly in individuals at higher risk of developing dementia (Subjective Cognitive Decline [SCD] and Mild Cognitive Impairment [MCI]). SCD is characterised by self-perceived cognitive decline without observable impairment on standardised cognitive tests, while MCI is defined by objective cognitive decline without impairment in functional ability. People meeting criteria for these conditions are a heterogenous group, with the aetiologies of symptoms spanning neurodegenerative, psychiatric and functional causes^3^.

Depression, characterised by persistent feelings of sadness and/or loss of interest in activities once enjoyed, has been identified as a dementia risk factor in the Lancet Commission on dementia prevention, intervention, and care, with preventing or treating depression estimated to prevent 3% of dementia cases^2^. Anxiety, characterised by persistent worry, has received less attention in dementia research. Nonetheless, several meta-analyses have examined associations between anxiety and dementia risk, with most finding that anxiety is associated with increased dementia risk^4,5^. There is a high prevalence of depression and anxiety symptoms in individuals with SCD and MCI^6^.

Physical inactivity has also been identified as a modifiable risk factor^2^ and has been linked with mental illness. A recent systematic review found that lower levels of sedentary behaviour were associated with lower anxiety symptoms^7^, and in cognitively healthy older adults, individuals with clinical depression had lower physical activity than those without^8^.

Given that individuals with SCD and MCI have an increased likelihood of experiencing depression and anxiety symptoms, and that physical inactivity is an established modifiable risk factor for dementia, understanding how step count relates to mental health in this population may offer valuable insights for early prevention. In people with SCD and MCI, the ability to engage in activities that raise step counts may be compromised by cognitive symptoms and by the physical health conditions that frequently co-occur with them. There is precedent for relationships differing in this population: for risk factors such as body mass index and blood pressure, midlife elevations predict increased dementia risk, yet low values in later life may instead mark prodromal neurodegeneration^9^. Associations observed in healthy older adults therefore cannot be assumed to hold in those at risk of dementia.

## OBJECTIVE

This study used data from APPLE-Tree, a randomised controlled trial of a group psychosocial lifestyle intervention of older adults with SCD or MCI, with objectively measured step count and depressive and anxiety symptoms assessed at baseline and at 12- and 24-month follow-up.

The primary objective was to investigate the cross-sectional associations between step count, depressive symptoms, and anxiety symptoms, which to our knowledge have not previously been examined in this population. The second aim was to examine whether self-perceived mobility mediated these associations. The APPLE-Tree intervention did not change step count in the full trial sample^10^; building on this, the third aim examined whether it changed step count specifically among those with clinical anxiety or depression at baseline. Finally, the fourth aim examined whether change in step count was associated with change in depressive or anxiety symptoms over time.

## METHODS

Data were drawn from the APPLE-Tree trial, a multi-site, randomised controlled trial of a remote multi-domain lifestyle intervention for older adults with SCD or MCI. Recruitment was conducted through multiple channels, including the English National Health Service, community partners, and social media platforms.

Eligible participants were aged 60 years or older and performed within educational- and age-adjusted normal ranges for MCI or SCD on the QuickMCI Screen^11^. Participants scoring <62 were classified as MCI. Those who scored 62+ and answered ‘yes’ to at least two of the three questions designed to detect SCD were classified as having SCD. Exclusion criteria included a diagnosis of dementia, a terminal condition, or harmful use of alcohol. Detailed eligibility criteria are described elsewhere^12^.

At baseline, 748 participants consented to participate. Two were excluded after baseline assessment (one mistakenly randomised twice, and one following review by the trial steering committee), resulting in 746 participants. Included and excluded participants were broadly similar across demographic and clinical characteristics (Supplementary Table S1).

The trial received ethical approval from the London (Camden and Kings Cross) Research Ethics Committee (Reference: 19/LO/0260) and UK Health Research Authority (HRA). All participants provided written informed consent.

### Intervention

Participants were randomised to usual care plus written dementia prevention information (control) or the 12-month APPLE-Tree intervention. The intervention has previously been described in detail^10^. It comprised ten fortnightly one-hour group video call sessions over six months, focused on lifestyle and behaviour change (e.g. diet, physical activity), with individual goal-setting calls after each session and informal group “tea breaks” between sessions. Over the subsequent six months, participants attended monthly group reunion sessions. Sessions were delivered remotely via Zoom owing to COVID-19.

### Measures

Assessments were collected at baseline, 12, and 24 months. Baseline measures were used for cross-sectional analyses, with these analyses repeated using 12- and 24-month data; step count, depressive and anxiety symptoms at all three timepoints were used for longitudinal analyses.

#### Step count

Participants wore consumer-grade wrist-worn wearables (Garmin Vivosmart 4) on their preferred wrist for 14 days at baseline, prior to undergoing any intervention. The watches did not have a visible read out of steps – to prevent this influencing activity levels, though participants were aware they were being worn for the primary purpose of counting steps. A valid wear day was one with at least 2 hours of wear, verified by concurrent heart-rate data. Average daily step count was computed from valid days; when worn beyond 14 days, the first 14 days were used.

#### Depressive and anxiety symptoms

These were assessed using the Hospital Anxiety and Depression Scale (HADS)^13^, comprising depression and anxiety subscales (each 0-21; higher indicates more symptoms; ≥8 indicates clinical levels^14^), which can be summed to a total score (0-42). The HADS has been validated and shown to have adequate psychometric properties^14^.

#### Covariates

Self-reported age, sex, ethnicity, education, and living arrangement were collected. Self-perceived mobility was measured by the EQ-5D-5L mobility item (five levels from “no problems” to “unable to walk”) and subjective health by the EQ-5D-5L visual analogue scale (0–100, higher suggests better perceived health). Primary support network size was measured using a seven-item questionnaire from the 1987 Health and Lifestyle Survey^15^ and categorised as no, moderate, or severe lack of social support. Cognitive status (MCI or SCD) reflected the presence or absence of objective cognitive impairment.

### Statistical Analysis

#### Cross-sectional analyses

Associations between step count, depressive and anxiety symptoms, and covariates were examined using Pearson’s correlation (continuous variables) and t-tests or ANOVA (categorical variables: sex, living arrangement, cognitive status, support network size).

Separate linear regressions for depressive and anxiety symptoms were run unadjusted (Model 1) and adjusted (Model 2) for age, sex, education, ethnicity, cognitive status, anxiety symptoms (for depression model) or depressive symptoms (for anxiety model), support network size, and living arrangement. Where an association was significant, interactions with sex and cognitive status were tested. Assumptions were checked before regression.

Sensitivity analyses (1) repeated the cross-sectional analyses using 12- and 24-month data; (2) used logistic regression for clinical versus non-clinical symptom levels; (3) examined HADS total score; (4) excluded outliers by removing participants with step counts exceeding 3 standard deviations above the mean; and (5) restricted to participants with ≥7 or ≥10 valid wear days.

The four-step mediation analysis by self-perceived mobility was tested using the Baron and Kenny framework^16^.

#### Longitudinal analyses

We examined whether the intervention’s effect on step count differed by baseline clinical anxiety or depression, using a three-level linear mixed-effects model as in the trial’s primary outcome paper^10^, with random effects for repeated measurements and therapy-group clustering in the intervention arm, and treatment group, baseline step count, site, time, and treatment × time as fixed effects; the baseline clinical group and its interactions with treatment and time tested whether the effect differed by group.

Separately, linear regression examined whether change in step count was associated with change in symptoms. Change scores (12- and 24-month follow-up minus baseline) were computed for step count and symptoms, and participants were categorised as having increased versus no change or decreased step count. The two groups were compared on follow-up symptom scores, adjusting for baseline symptom score and baseline step count. Models were repeated adjusting for trial arm and with step count change modelled continuously.

All analyses were performed in R (version 4.2.2). Standardised beta coefficients are reported.

## FINDINGS

### Cross-sectional analyses

Participants wore the device for an average of 13 days (SD = 2.3); adherence was high (96.3% wore it ≥7 days, 73.6% the full 14 days). With respect to mental health, 422 participants (67.1%) had non-clinical levels of depression or anxiety, 26 (4.1%) met clinical thresholds for depression alone, 109 (17.3%) for anxiety alone, and 72 (11.4%) for both.

Higher step count was associated with fewer depressive symptoms (r = −0.15, p < 0.001), more anxiety symptoms (r = 0.09, p = 0.02), better subjective health (r = 0.28, p < 0.001), and younger age (r = −0.36, p < 0.001). Depressive and anxiety symptoms were strongly correlated (r = 0.60, p < 0.001), and both were associated with worse subjective health (depression: r = −0.47, p < 0.001; anxiety: r = −0.30, p < 0.001). Anxiety symptoms were additionally associated with younger age (r = −0.26, p < 0.001).

Group comparisons revealed several differences. Women reported higher anxiety symptoms than men (p < 0.001), with no difference in depressive symptoms (p = 0.11) or step count (p = 0.88). Participants living alone took fewer daily steps (p = 0.001) and reported more depressive symptoms (p = 0.005) than those living with others, with no difference in anxiety (p = 0.43). Participants with MCI took fewer steps than those with SCD (p < 0.001) but did not differ in depressive (p = 0.08) or anxiety (p = 0.34) symptoms.

#### Assumptions of regression models

Assumptions of linearity and independence were met. Homoscedasticity and normality were mildly violated; however, as linear regression is robust to such departures^17^, we proceeded with linear models for all analyses.

#### Association between step count and depressive symptoms

Higher step count was associated with lower depressive symptoms in the unadjusted (Model 1: β = −0.15, 95% CI: −0.23 to −0.07, p < 0.001) and adjusted (Model 2: β = −0.11, 95% CI: −0.17 to −0.05, p < 0.001; Figure 1) models. Excluding participants with step counts exceeding 3 standard deviations from the mean (n = 5 excluded) did not change the results (Model 2: β = −0.13, 95% CI: −0.19 to −0.07, p < 0.001). This association was consistent in cross-sectional analyses using 12- and 24-month data (Supplementary Table S2).

**Figure 1.**
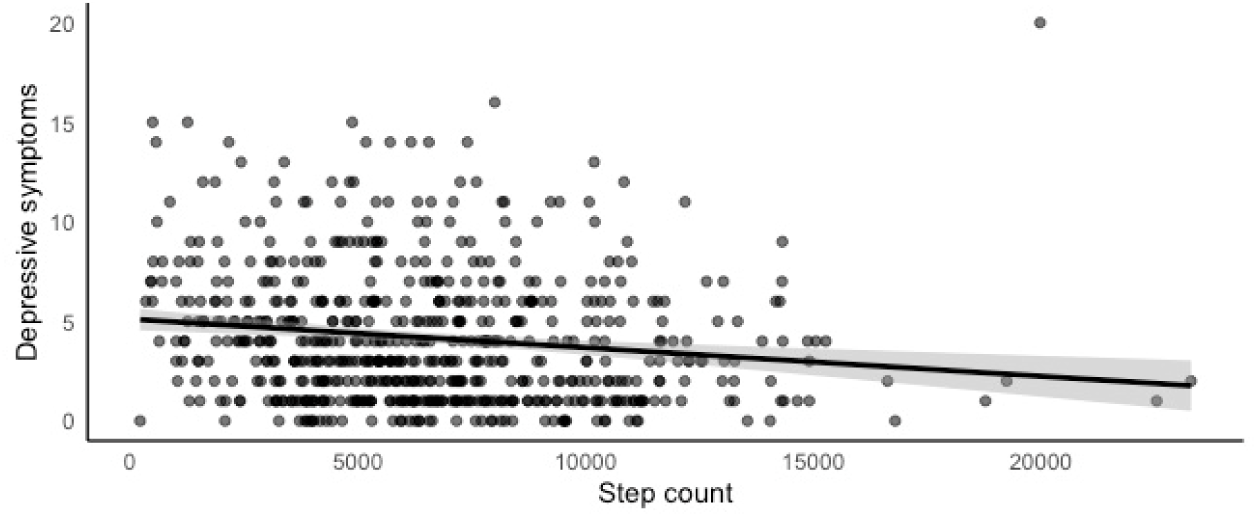
Association between step count and depressive symptoms Association between step count and depressive symptoms. Higher step count was associated with fewer depressive symptoms (adjusted β = −0.11, 95% CI −0.17 to −0.05, p < 0.001). Model adjusted for age, sex, education, ethnicity, cognitive status, subjective health, primary support network size, living arrangement, and anxiety symptoms.

**Table 1.** Sample characteristics.

| **Variable** | **Mean or N** | **SD or %** |
| --- | --- | --- |
| **Demographics** |  |  |
| Age | 74.21 | 6.87 |
| Sex |  |  |
| Female | 296 | 47.06 % |
| Male | 333 | 52.94% |
| Ethnicity |  |  |
| Asian | 40 | 6.36% |
| Black | 7 | 1.11% |
| Mixed | 14 | 2.23% |
| Other | 6 | 0.95% |
| White UK | 511 | 81.24% |
| White Other | 51 | 8.11% |
| Marital status |  |  |
| Single | 40 | 6.36% |
| Married/civil partnership | 400 | 63.59% |
| Living with partner | 26 | 4.13% |
| Widowed | 87 | 13.83% |
| Divorced | 75 | 11.92% |
| Unable to specify | 1 | 0.16% |
| Highest level of education |  |  |
| No Education | 2 | 0.32% |
| Primary | 8 | 1.27% |
| Secondary (e.g. O level; GCSE) | 137 | 21.78% |
| Further (e.g. A level; BTEC; NVQ) | 171 | 27.19% |
| Degree | 171 | 27.19% |
| Postgraduate | 132 | 20.99% |
| Other | 7 | 1.11% |
| Unable to specify | 1 | 0.16% |
| Employment |  |  |
| Full time employment | 30 | 4.77% |
| Part time employment | 48 | 7.63% |
| Retired | 510 | 81.08% |
| Unemployed/unable to work | 16 | 2.54% |
| Other | 24 | 3.82% |
| Unable to specify | 1 | 0.16% |
| Living situation |  |  |
| Live alone | 166 | 26.39% |
| Live with others | 463 | 73.61% |
| **Psychiatric symptoms** |  |  |
| Depressive symptoms | 4.16 | 3.37 |
| Categorical Depression |  |  |
| Clinical symptoms ($\geq 8$) | 98 | 15.58% |
| Non-clinical symptoms ($< 8$) | 531 | 84.42% |
| Anxiety symptoms | 5.57 | 4.04 |
| Categorical Anxiety |  |  |
| Clinical symptoms ($\geq 8$) | 181 | 28.79% |
| Non-clinical symptoms ($< 8$) | 448 | 71.22% |
| HADS total score | 9.73 | 6.63 |
| **Step count** |  |  |
| Mean (SD) | 6585.81 | 3515.78 |
| Median (range) | 6242 | 217-23296 |
| Number of days worn | 12.99 | 2.33 |
| **Cognitive status** |  |  |
| SCD | 379 | 60.25% |
| MCI | 249 | 39.59% |
| Missing | 1 | 0.16% |
| **Primary support network size** | 19.80 | 2.28 |
| No lack of social support | 152 | 24.17% |
| Moderate lack of social support | 395 | 62.80% |
| Severe lack of social support | 81 | 12.88% |
| Missing data | 1 | 0.16% |
| **Self-perceived mobility problems<sup>1</sup>** |  |  |
| No problems in walking | 390 | 62.00% |
| Slight problems in walking | 135 | 21.46% |
| Moderate problems in walking | 77 | 12.24% |
| Severe problems in walking | 27 | 4.29% |
| **Subjective health (n = 328)<sup>2</sup>** | 76.40 | 16.84 |

Abbreviations: HADS; Hospital Anxiety and Depression Scale; SCD; Subjective Cognitive Decline; SD, standard deviation; MCI, Mild Cognitive Impairment
<sup>1</sup>Measured using EuroQoL EQ-5D
<sup>2</sup>Participants were asked to rate how good or bad their health is today, on a scale from 0 (the worst health you can imagine) to 100 (the best health you can imagine)

No statistically significant interactions were observed between step count and sex or cognitive status on depressive symptoms in both the unadjusted and adjusted models (Supplementary Table S3).

Sensitivity analyses revealed that higher step count was associated with a lower likelihood of having clinical levels of depressive symptoms in unadjusted and adjusted models (Supplementary Table S3). All results remained unchanged when using data from participants with ≥7 or ≥10 valid wear days (Supplementary Table S4).

#### Association between step count and anxiety symptoms

Higher step count was associated with more anxiety symptoms in both models (Model 1: β = 0.09, 95% CI: 0.01 to 0.17, p = 0.02; Model 2: β = 0.12, 95% CI: 0.06 to 0.19, p = 0.003; Figure 2). Excluding participants with step counts exceeding 3 standard deviations from the mean (n = 5 excluded) did not change the results (Model 2: β = 0.13, 95% CI: 0.07 to 0.20, p = 0.001). In cross-sectional analyses using 12- and 24-month data, higher step count remained associated with more anxiety symptoms in the adjusted models at both timepoints, although the unadjusted associations were not statistically significant (Supplementary Table S2).

**Figure 2.**
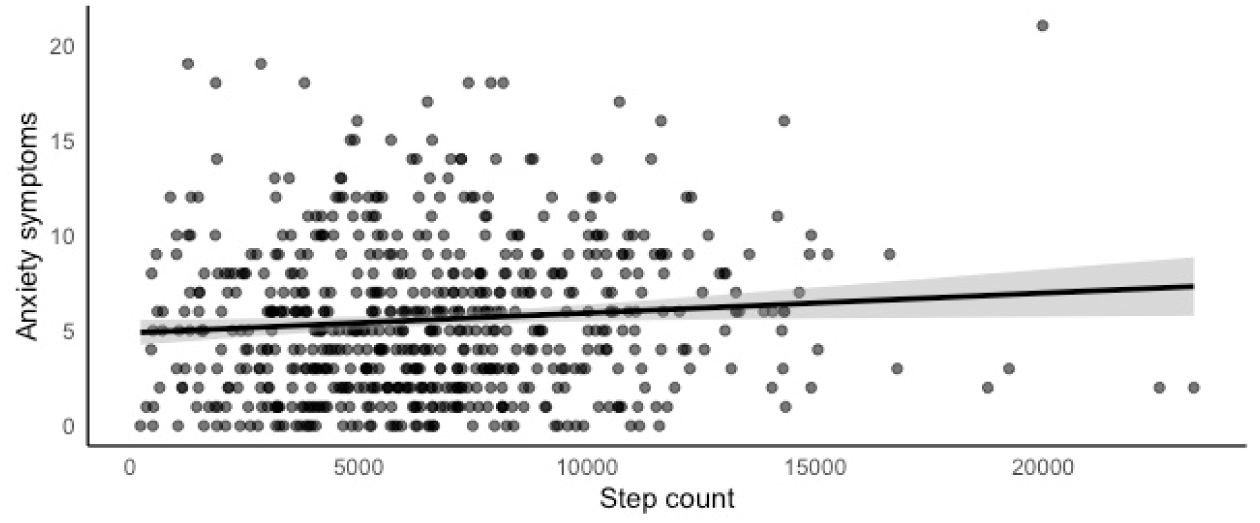
Association between step count and anxiety symptoms Association between step count and anxiety symptoms. Higher step count was associated with more anxiety symptoms (adjusted β = 0.12, 95% CI 0.06 to 0.19, p = 0.003). Model adjusted for age, sex, education, ethnicity, cognitive status, subjective health, primary support network size, living arrangement, and depressive symptoms.

No statistically significant interactions were observed between step count and sex or cognitive status on anxiety symptoms in both the unadjusted and adjusted models (Supplementary Table S5).

Sensitivity analyses revealed that higher step count was associated with a higher likelihood of having clinical (vs. non-clinical) levels of anxiety symptoms in the adjusted model, but not in the unadjusted model (Supplementary Table S5). Findings from sensitivity analyses using data from participants with ≥7 or ≥10 valid wear days were consistent with the main results in the adjusted model but not the unadjusted model (Supplementary Table S6).

#### Association between step count and HADS total score

No associations were observed between step count and total HADS score (Model 1: β = −0.02, 95% CI: −0.10 to 0.06, p = 0.61; Model 2: β = 0.02, 95% CI: −0.05 to 0.09, p = 0.61). Sensitivity analyses using data from participants with ≥7 or ≥10 valid wear days showed findings consistent with the main results (Supplementary Table S7).

#### Mediation analyses

Higher step count was associated with lower depressive symptoms (β = −0.15, 95% CI −0.23 to −0.07, p < 0.001; Figure 3a) and with lower self-perceived mobility problems (β = −0.35, 95% CI −0.41 to −0.28, p < 0.001), which were in turn associated with more depressive symptoms after controlling for step count (β = 0.44, 95% CI 0.35 to 0.54, p < 0.001). After adjusting for self-perceived mobility, step count was no longer associated with depressive symptoms (β = 0.004, 95% CI −0.08 to 0.08, p = 0.92), indicating full mediation.

**Figure 3a.**
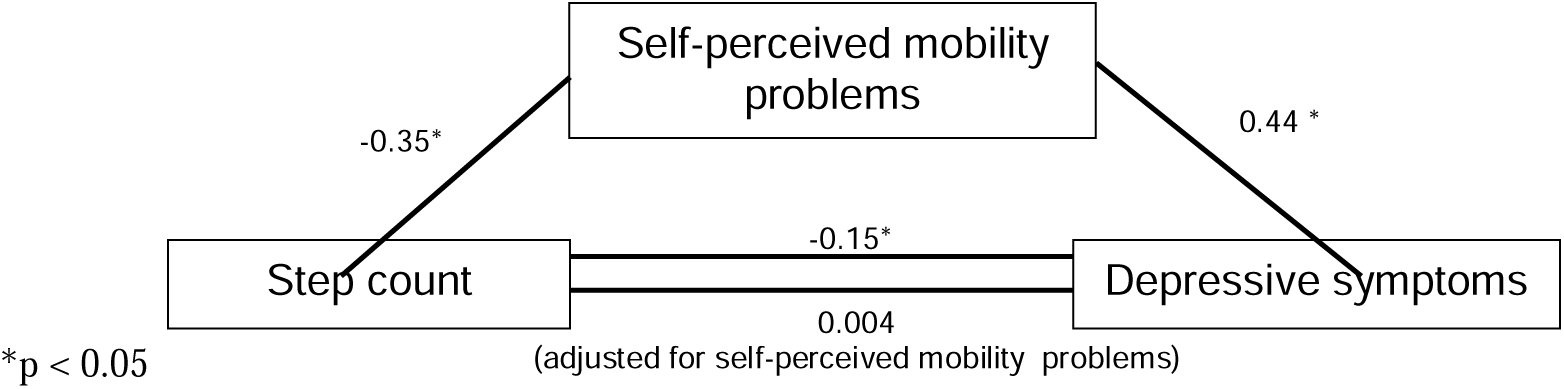
Standardised regression coefficients for the association between step count and depressive symptoms as mediated by self-perceived mobility problems.

For anxiety, higher step count was associated with more anxiety symptoms (β = 0.09, 95% CI 0.01 to 0.17, p = 0.02; Figure 3b) and with lower self-perceived mobility problems (β = −0.35, 95% CI −0.41 to −0.28, p < 0.001), which were associated with more anxiety symptoms after controlling for step count (β = 0.25, 95% CI 0.15 to 0.35, p < 0.001). The step count-anxiety association remained significant after adjusting for self-perceived mobility (β = 0.18, 95% CI 0.09 to 0.26, p < 0.001), indicating no mediation.

**Figure 3b.**
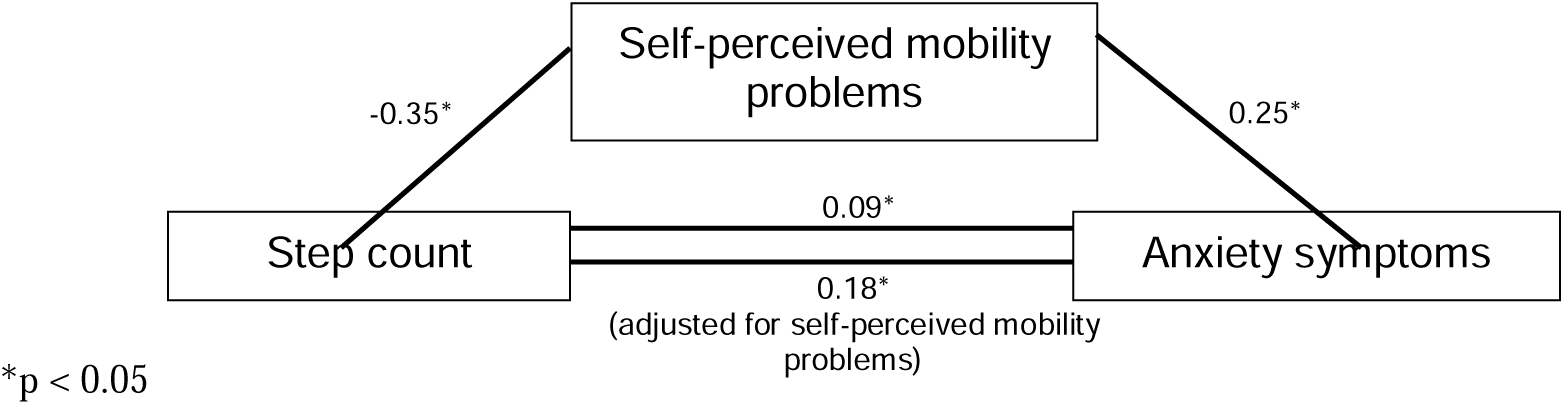
Standardised regression coefficients for the association between step count and anxiety symptoms as mediated by self-perceived mobility problems.

### Longitudinal analyses

#### Effect of intervention on step count

Step count data were available for 430 participants at 12 months and 240 at 24 months. Among these, 124 of the 181 participants with baseline clinical anxiety and 60 of the 98 with baseline clinical depression had step count data at 12 months (66 and 35, respectively, at 24 months). There was no significant effect of the intervention on step count at 12 or 24 months in participants with clinical anxiety or depression (Supplementary Table S8), and no evidence that its effect differed between clinical and non-clinical participants (interaction term: anxiety p = 0.25; depression p = 0.86).

#### Change in step count and change in symptoms

Change in step count from baseline was not associated with change in depressive or anxiety symptoms. At 12 months (376 participants with step count and symptom data at both baseline and 12 months), change in depressive symptoms did not differ between participants whose step count increased and those with no change or a decrease (adjusted difference = −0.19, 95% CI −0.65 to 0.28, p = 0.43); nor did change in anxiety symptoms (−0.10, 95% CI −0.63 to 0.43, p = 0.72). Findings were similar at 24 months (215 participants with step count and symptom data at both baseline and 24 months; depression: −0.50, 95% CI −1.13 to 0.13, p = 0.12; anxiety: 0.24, 95% CI −0.50 to 0.99, p = 0.52). Results were consistent when adjusting for trial arm and when step count change was modelled continuously.

## DISCUSSION

This study examined associations between step count, depressive symptoms, and anxiety symptoms in older adults at risk of dementia. In cross-sectional analyses, higher step counts were associated with fewer depressive symptoms but, contrary to expectations, with more anxiety symptoms. Associations did not differ by cognitive status or sex, and no association was found with total HADS score. Self-perceived mobility problems mediated the association between step count and depressive symptoms, but not anxiety symptoms. In longitudinal analyses, the intervention did not change step count in those with clinical anxiety or depression, and change in step count was not associated with change in depressive or anxiety symptoms.

### Cross-sectional findings

#### Step count and depressive symptoms

Higher daily step count has been consistently associated with lower depressive symptoms in adult^18^ and older adult populations^19^. A meta-analysis of objectively measured step count found greater daily step count was associated with fewer depressive symptoms, with associations significant in older adults and in studies using wrist-worn devices^20^. The present study extends this evidence to older adults with SCD or MCI, finding that higher step count was associated with fewer depressive symptoms after adjusting covariates.

Several mechanisms may underlie this association. Physical activity has been shown to promote neuroplasticity through increased production of brain-derived neurotrophic factor, which supports neuronal survival, growth, and synaptic plasticity in brain regions involved in mood regulation^21^. Psychological mechanisms may also contribute. For example, physical activity has been found to reduce depressive symptoms through enhanced physical activity self-efficacy^22^. However, while objectively measured step count reflects actual behaviour, it does not capture how individuals perceive their own mobility – perceptions that may influence mental health independently of objective activity levels^23^. The present study found that self-perceived mobility problems mediated the association between step count and depressive symptoms, suggesting that individuals with higher step counts may experience fewer depressive symptoms partly through more positive perceptions of their ability to be mobile. This is consistent with previous work showing self-perceived mobility problems mediates the relationship between sedentary behaviour and depressive symptoms across age groups and in older adults^23^. To our knowledge, this is the first study to examine self-perceived mobility problems as a mediator between step count and depressive symptoms in this population.

#### Step count and anxiety symptoms

This study found that higher step count was associated with more anxiety symptoms and an increased likelihood of clinical anxiety after adjustment for relevant covariates. This positive association contrasts with existing literature, which typically reports negative associations between physical activity and anxiety. For example, greater physical activity measured using wrist-worn wearables were associated with a lower risk of anxiety in a large prospective cohort of middle-aged and older adults^24^. However, not all findings align: one study found no association between step count and anxiety symptoms in middle-aged adults^25^, and a large study of adults identified a U-shaped association, with physical activity above 6,000 MET-minutes (metabolic equivalent of task) per week showing no reduced risk of clinical anxiety^26^.

One explanation for the positive association observed here is that high step count may reflect restlessness (characterised by difficulty staying still), which is commonly experienced by people with anxiety^27^. In such cases, increased step count may reflect anxiety-driven movement rather than intentional health-promoting activity. This may have been accentuated by the study context: data were collected during the COVID-19 pandemic, when restrictions on outdoor activities and social distancing limited opportunities for structured, purposeful walking and elevated anxiety in older adults^28^. Higher step counts may have reflected anxious or restless indoor movement rather than health-promoting outdoor walking. However, the positive association persisted in cross-sectional analyses at 12 and 24 months, by which point pandemic restrictions had largely eased, suggesting that the association is unlikely to be explained by the pandemic context alone.

Higher step count may also reflect health anxiety. People with cognitive concerns are a heterogeneous group, with some individuals anxious about developing dementia who may therefore engage intensively in recommended risk-reduction behaviours, including increasing physical activity. A systematic review found that use of fitness and diet trackers was associated with disordered eating and excessive exercise, suggesting self-monitoring of health behaviours may heighten anxious preoccupation^29^. Similarly, tracking step counts may sustain anxiety, such that higher step count and greater anxiety reinforce one another.

### Longitudinal findings

The APPLE-Tree intervention did not change step count in the full trial sample^10^, and we found no evidence that it changed step count even among those with clinical anxiety or depression at baseline. Although we observed cross-sectional associations between step count and both anxiety and depressive symptoms, change in step count was not associated with change in symptoms over time. Step count may therefore be understood as a correlate or marker of depressive and anxiety symptoms in this population – potentially useful for identifying co-occurring symptoms – rather than a target that, if increased, would itself reduce them. Whether step count and symptoms are causally related, and in which direction, cannot be determined from these data.

### Strengths and limitations

This study has several strengths. It addresses gaps in the literature by using objectively measured step counts in older adults with SCD or MCI, employs a comprehensive statistical approach with carefully considered covariates, and uses consumer-grade wrist-worn wearables obscured with black stickers to minimise reactivity. Consumer-grade devices also enhance ecological validity given their widespread availability. Repeated assessments at 12 and 24 months also allowed us to examine change over time.”

Several limitations should be noted. First, this was a trial sample rather than an epidemiological one. Participants who volunteer for a dementia-prevention randomised controlled trial are likely to differ from the wider population of people with cognitive concerns – for example, in being more motivated, or engaged with lifestyle change – which may limit the generalisability of these findings. Second, whether wearables were worn on the dominant or non-dominant wrist was not recorded; since wrist-worn devices measure step count based on wrist and arm movements, dominant-hand wear may have inflated step counts. Third, the APPLE-Tree protocol defined a valid day as at least two hours of wear time, which maximises inclusion but may underestimate typical activity for participants with shorter wear durations. Fourth, although we were able to examine longitudinal change, these analyses were limited by the small number of participants with clinical anxiety or depression at follow-up.

### Clinical implications and future research

To our knowledge, this is the first study to examine objectively measured step counts in relation to depressive and anxiety symptoms in older adults with SCD or MCI. Future research is needed to replicate these findings and clarify why the relationship between step count and anxiety in this population differs from that typically observed in cognitively healthy older adults, or whether this is context-specific (e.g., data collection during the COVID-19 pandemic). Future work should also determine whether the observed association reflects anxiety-related movement such as restlessness; detailed examination of the context and intentionality of movement (e.g., purposeful versus unstructured walking) will be important.

Although depression and anxiety symptoms are strongly correlated, this study demonstrates that they have opposing associations with step counts, highlighting the importance of assessing them separately. Evidence from intervention studies suggests that physical activity, including walking, can reduce depressive and anxiety symptoms in healthy older adults^30^. However, we found no evidence that the APPLE-Tree intervention changed step count, or that change in step count was associated with change in depressive and anxiety symptoms in these older adults with SCD or MCI. Step count may therefore be more useful as a marker of co-occurring symptoms than as an intervention target in people with SCD or MCI. Future research is needed to clarify the underlying mechanisms, and larger longitudinal studies are required, before walking interventions can be confidently recommended for anxiety or depression in this group.

## Conclusion

This study investigated associations between step count, depressive symptoms, and anxiety symptoms in older adults with SCD or MCI. Consistent with existing literature, higher step count was associated with lower depressive symptoms. Contrary to most existing evidence, higher step count was associated with more anxiety symptoms – an association that may reflect restlessness, although future research is needed to validate this interpretation. The associations may be driven by different mechanisms: self-perceived mobility problems mediated the relationship with depressive symptoms but not anxiety symptoms. In longitudinal analyses, neither the intervention nor within-person change in step count was associated with change in symptoms. Future studies should distinguish health-promoting walking from anxiety-driven movement to better understand the mechanisms linking step count with anxiety in older adults at risk of dementia.

## Data Availability

All data produced in the present study are available upon reasonable request to the authors.

