## Supplementary Materials for "Differential association of step count with depressive and anxiety symptoms in older adults at risk of dementia"

**Table S1.** Sample characteristics

| **Variable** | **Sample in this study**  **(n = 629)** | **Total APPLE-tree sample**  **(n = 746)** |
| --- | --- | --- |
| **Demographics** |  |  |
| **Age** | 74.21 (6.87) | 74.45 (6.93) |
| **Sex** |  |  |
| Female | 296 (47.06%) | 351 (47.05%) |
| Male | 333 (52.94%) | 392 (52.54%) |
| **Ethnicity** |  |  |
| Asian | 40 (6.36%) | 50 (6.74%) |
| Black | 7 (1.11%) | 13 (1.75%) |
| Mixed | 14 (2.23%) | 16 (2.15%) |
| Other | 6 (0.95%) | 7 (0.94%) |
| White UK | 511 (81.24%) | 599 (80.73%) |
| White Other | 51 (8.11%) | 57 (7.68%) |
| **Marital status** |  |  |
| Single | 40 (6.36%) | 46 (6.17%) |
| Married/civil partnership | 400 (63.59%) | 464 (62.20%) |
| Living with partner | 26 (4.13%) | 30 (4.02%) |
| Widowed | 87 (13.83%) | 108 (14.48%) |
| Divorced | 75 (11.92%) | 92 (12.33%) |
| Unable to specify | 1 (0.16%) | 2 (0.27%) |
| **Highest level of education** |  |  |
| No Education | 2 (0.32%) | 2 (0.27%) |
| Primary | 8 (1.27%) | 10 (1.34%) |
| Secondary (e.g. O level; GCSE) | 137 (21.78%) | 167 (22.39%) |
| Further (e.g. A level; BTEC; NVQ) | 171 (27.19%) | 199 (26.68%) |
| Degree | 171 (27.19%) | 204 (27.35%) |
| Postgraduate | 132 (20.99%) | 147 (19.71%) |
| Other | 7 (1.11%) | 11 (1.47%) |
| Unable to specify | 1 (0.16%) | 2 (0.27%) |
| **Living situation** |  |  |
| Live alone | 166 (26.39%) | 198 (26.54%) |
| Live with others | 463 (73.61%) | 538 (72.12%) |
| **Psychiatric symptoms** |  |  |
| Depressive symptoms | 4.16 (3.37) | 4.18 (3.43) |
| Anxiety symptoms | 5.57 (4.04) | 5.57 (4.07) |
| **Step count** | 6585.81 (3515.78) | 6602.70 (3506.78) |
| **Cognitive status**^1^ |  |  |
| SCD | 379 (60.25%) | 436 (58.45%) |
| MCI | 249 (39.59%) | 309 (41.24%) |
| **Loneliness** | 4.30 (1.67) | 4.23 (1.79) |
| **Primary support network size**^1^ | 19.80 (2.28) | 19.80 (2.28) |
| **Subjective health^1^** | 76.40 (16.84) | 76.00 (17.00) |

Abbreviations: HADS; Hospital Anxiety and Depression Scale; MCI, Mild Cognitive Impairment; SCD; Subjective Cognitive Decline; SD, standard deviation

^1^One participant with missing data

**Table S2.** Cross-sectional association between step count and depressive and anxiety symptoms at 12- and 24-month follow-up

|  | **β (95% CI)** | **p-value** |
| --- | --- | --- |
| **Depressive symptoms** |  |  |
| **12 months (n = 413 unadjusted; 375 adjusted)** | | |
| Model 1 | -0.19 (-0.28 to -0.10) | <0.001 |
| Model 2 | -0.14 (-0.22 to -0.06) | <0.001 |
| **24 months (n = 240 unadjusted; 215 adjusted)** | | |
| Model 1 | -0.27 (-0.39 to -0.15) | <0.001 |
| Model 2 | -0.24 (-0.34 to -0.13) | <0.001 |
| **Anxiety symptoms** |  |  |
| **12 months (n = 413 unadjusted; 375 adjusted)** | | |
| Model 1 | 0.05 (-0.04 to 0.14) | 0.29 |
| Model 2 | 0.12 (0.04 to 0.20) | 0.005 |
| **24 months (n = 240 unadjusted; 215 adjusted)** | | |
| Model 1 | 0.05 (-0.07 to 0.18) | 0.41 |
| Model 2 | 0.19 (0.07 to 0.30) | 0.002 |

Model 1: unadjusted

Model 2: adjusted for age, sex, education, and ethnicity, cognitive impairment level, subjective health, primary support network size and living arrangement

**Table S3.** Depression model interaction and sensitivity analyses

|  | **β (95% CI)** | **p-value** |
| --- | --- | --- |
| **Interaction with sex (reference group: women)** | | |
| Model 1 | 0.02 (-0.13 to 0.18) | 0.79 |
| Model 2 | 0.03 (-0.08 to 0.15) | 0.59 |
| **Interaction with cognitive impairment (reference group: subjective cognitive decline)** | | |
| Model 1 | 0.06 (-0.10 to 0.22) | 0.46 |
| Model 2 | 0.07 (-0.05 to 0.18) | 0.27 |
|  | **ORs (95% CI)** | **p-value** |
| **Sensitivity analyses (clinical vs. non-clinical depression)** | | |
| Model 1 | 0.68 (0.53 to 0.86) | 0.002 |
| Model 2 | 0.55 (0.38 to 0.79) | 0.001 |

OR, Odds Ratio; CI, Confidence Interval; SD

Model 1: unadjusted

Model 2: adjusted for age, sex, education, and ethnicity, cognitive impairment level, anxiety symptoms, subjective health, primary support network size and living arrangement

**Table S4.** Sensitivity analyses: Association between step count and depressive symptoms among participants with ≥7 days or ≥10 days of wearable data

|  | **β (95% CI)** | **p-value** |
| --- | --- | --- |
| **≥7 days (n = 606)** | | |
| Model 1 | -0.19 (-0.27 to -0.11) | <0.001 |
| Model 2 | -0.13 (-0.20 to -0.07) | <0.001 |
| **≥10 days (n = 583)** | | |
| Model 1 | -0.20 (-0.27 to -0.11) | <0.001 |
| Model 2 | -0.13 (-0.20 to -0.07) | <0.001 |

Model 1: unadjusted

Model 2: adjusted for age, sex, education, and ethnicity, cognitive impairment level, anxiety symptoms, subjective health, primary support network size and living arrangement

**Table S5.** Anxiety model interaction and sensitivity analyses

|  | **β (95% CI)** | **p-value** |
| --- | --- | --- |
| **Interaction with sex (reference group: women)** | | |
| Model 1 | -0.04 (-0.19 to 0.11) | 0.62 |
| Model 2 | -0.04 (-0.16 to 0.07) | 0.47 |
| **Interaction with cognitive impairment (reference group: subjective cognitive decline)** | | |
| Model 1 | -0.002 (-0.16 to 0.16) | 0.98 |
| Model 2 | -0.04 (-0.16 to 0.09) | 0.55 |
|  | **ORs (95% CI)** | **p-value** |
| **Sensitivity analyses (clinical and non-clinical anxiety)** | | |
| Model 1 | 1.18 (0.99 to 1.40) | 0.06 |
| Model 2 | 1.40 (1.09 to 1.80) | 0.01 |

OR, Odds Ratio; CI, Confidence Interval; SD

Model 1: unadjusted

Model 2: adjusted for age, sex, education, and ethnicity, cognitive impairment level, depressive symptoms, subjective health, primary support network size and living arrangement

**Table S6**. Sensitivity analyses: Association between step count and anxiety symptoms among participants with ≥7 days or ≥10 days of wearable data

|  | **β (95% CI)** | **p-value** |
| --- | --- | --- |
| **≥7 days (n = 606)** | | |
| Model 1 | 0.01 (-0.01 to 0.15) | 0.09 |
| Model 2 | 0.11 (0.02 to 0.15) | 0.01 |
| **≥10 days (n = 583)** | | |
| Model 1 | 0.06 (-0.02 to 0.14) | 0.13 |
| Model 2 | 0.12 (0.05 to 0.19) | 0.001 |

Model 1: unadjusted

Model 2: adjusted for age, sex, education, and ethnicity, cognitive impairment level, depressive symptoms, subjective health, primary support network size and living arrangement

**Table S7.** Sensitivity analyses: Association between step count and total HADS among participants with ≥7 days or ≥10 days of wearable data

|  | **β (95% CI)** | **p-value** |
| --- | --- | --- |
| **≥7 days (n = 606)** | | |
| Model 1 | -0.05 (-0.13 to 0.03) | 0.19 |
| Model 2 | -0.01 (-0.09 to 0.07) | 0.80 |
| **≥10 days (n = 583)** | | |
| Model 1 | -0.06 (-0.14 to 0.02) | 0.14 |
| Model 2 | -0.02 (-0.01 to 0.06) | 0.59 |

Model 1: unadjusted

Model 2: adjusted for age, sex, education, and ethnicity, cognitive impairment level, subjective health, primary support network size and living arrangement

**Table S8**. Effect of the intervention on step count at 12 and 24 months, stratified by baseline clinical anxiety and clinical depression

|  | **Control n** | **Control Mean (SD)** | **Intervention n** | **Intervention Mean (SD)** | **Adjusted mean difference (95% CI), p** |
| --- | --- | --- | --- | --- | --- |
| **Clinical anxiety at baseline** | | | | | |
| 12 months | 58 | 7133.5 (3106.9) | 66 | 6715.7 (3253.3) | −237.5 (−1079.4 to 604.0), p = 0.58 |
| 24 months | 31 | 7185.2 (3907.8) | 35 | 6320.9 (3138.2) | −358.4 (−1438.6 to 721.8), p = 0.51 |
| **Clinical depression at baseline** | | | | | |
| 12 months | 29 | 5633.9 (3344.0) | 31 | 5430.2 (3332.0) | −119.9 (−1313.0 to 1073.0), p = 0.84 |
| 24 months | 17 | 4588.4 (3905.3) | 18 | 5004.6 (2484.7) | +161.7 (−1344.0 to 1668.0), p = 0.83 |
